# Emerging Cryptococcosis cases in Himalayan & Sub-Himalayan regions of India

**DOI:** 10.1101/2025.11.05.25339595

**Authors:** Arpita Subhashree, Rajat Sharma, Amber Prasad, Sowjanya Perumalla, Prasan Kumar Panda

## Abstract

**Background:** Cryptococcosis is a life-threatening opportunistic fungal infection, predominantly affecting immunocompromised individuals such as people living with HIV (PLHIV), but increasingly recognized among non-HIV patients with chronic comorbidities. Despite being a major contributor to meningitis-related mortality, there is limited regional data from North India, especially the Himalayan belt.

**Aim:** To describe the clinical, laboratory, and epidemiological profile of microbiologically confirmed cryptococcosis in a tertiary care center in Northern India and identify predictors of morbidity and mortality.

**Methods:** A retrospective observational study was conducted at AIIMS Rishikesh from January 2018 to December 2024. Adult patients (>15 years) with confirmed cryptococcosis were included. Data were extracted using structured REDCap forms and analyzed for demographics, risk factors, clinical presentation, treatment, and outcomes. Subgroup analyses and logistic regression were performed.

**Results:** Among 33 cases, 66.7% were HIV-positive. Fever (90.9%), headache (78.8%), and altered sensorium (54.5%) were the most common symptoms. Amphotericin B was used in 90.9% of patients, while flucytosine and fluconazole were administered in 45.5% and 66.7%, respectively. In-hospital mortality was 21.2%; however, with the inclusion of 1-year follow-up, mortality went up to 51.5%. Uncontrolled diabetes mellitus was significantly associated with mortality (p=0.019). Amphotericin B use was significantly lower among Himalayan patients (p=0.030), correlating with a trend toward higher mortality.

**Conclusion:** Cryptococcosis is associated with high morbidity and mortality in both HIV and non-HIV patients. Prompt diagnosis and equitable antifungal access, especially in Himalayan regions, are critical to improving outcomes.

## Introduction

Cryptococcosis is a globally prevalent opportunistic fungal infection caused predominantly by two encapsulated yeast species: *Cryptococcus neoformans* and *Cryptococcus gattii*, members of the *Tremellomycetes* class of Basidiomycota. These fungi are widely distributed in the environment, particularly in soil contaminated with bird droppings or decaying wood. Inhalation of airborne spores especially in immunocompromised is the principal route of transmission, leading to pulmonary infection, which may disseminate hematogenously, particularly to the central nervous system (CNS) [1]. The clinical spectrum of cryptococcosis ranges from asymptomatic colonization and mild pulmonary involvement to life-threatening meningoencephalitis, especially in immunocompromised individuals. It is recognized as one of the most important fungal infections in people living with HIV (PLHIV), especially in low- and middle-income countries, where cryptococcal meningitis accounts for a substantial proportion of AIDS-related deaths [2] [3]. Globally, an estimated 220,000 new cases of cryptococcal meningitis occur annually among PLHIV, resulting in nearly 180,000 deaths [4].

While *C. neoformans* predominantly affects immunocompromised hosts, including PLHIV, organ transplant recipients, and those receiving corticosteroids or immunosuppressants, *C. gattii* has a wider host range, often causing disease in apparently immunocompetent individuals [5]. Non-*neoformans* species such as *C. albidus* and *C. laurentii* have also emerged as rare but relevant pathogens in recent years [6].

In India, cryptococcosis is increasingly recognized as a major opportunistic infection, particularly in the context of the HIV epidemic. However, there remains limited literature on the disease burden and clinical characteristics of cryptococcosis from Northern India, particularly the Himalayan and sub-Himalayan regions, which pose unique healthcare challenges due to topographical inaccessibility and delayed diagnosis.

The CNS is the most commonly affected organ in disseminated cryptococcosis, presenting as subacute or chronic meningitis with symptoms such as headache, fever, vomiting, altered sensorium, neck stiffness, and cranial nerve palsies. These symptoms often mimic other causes of chronic meningitis such as tuberculosis, leading to misdiagnosis and delayed treatment [7]. Pulmonary cryptococcosis, though less dramatic, can manifest with cough, pleuritic chest pain, and radiographic findings ranging from nodules to consolidation or cavitation [8].

The management of cryptococcosis has advanced with the availability of rapid diagnostics like cryptococcal latex agglutination or cryptococcal antigen (CrAg) lateral flow assays and potent antifungal agents such as amphotericin B, flucytosine, and fluconazole. However, despite these advancements, the mortality associated with cryptococcal meningitis remains high, particularly in patients with delayed diagnosis or inadequate immune reconstitution [9].

This study aims to fill the knowledge gap in the regional and immunological variation of cryptococcosis presentations and outcomes in Northern India. Through a retrospective analysis of microbiologically confirmed cases at a tertiary care center in Uttarakhand over a 7-year period, we seek to delineate the demographic, clinical, and laboratory profile of cryptococcosis and identify predictors of morbidity and mortality in this underserved population.

## Materials and methods

### Study Design

A hospital-based, retrospective observational study was conducted on adult cryptococcosis cases (>15 years) admitted to All India Institute of Medical Sciences (AIIMS) Rishikesh between January 2018 and December 2024.

### Setting

The study was performed in the Department of General Medicine of AIIMS Rishikesh, a tertiary care referral hospital in Uttarakhand, Northern India. The hospital receives referrals from both Himalayan and sub-Himalayan zones. Himalayan region, was defined as areas located at an altitude above 1120 feet from sea level, including patients residing in all districts of Uttarakhand, except Haridwar district. Non-Himalayan included patients residing in lower-altitude adjoining areas such as Uttar Pradesh, Haryana, and Haridwar district of Uttarakhand.

### Participants

Cases were identified through microbiological records. Inclusion criteria comprised patients with laboratory-confirmed cryptococcosis (positive India ink, cryptococcal antigen test, or culture) admitted under Internal Medicine. Patients admitted to other departments without infectious disease consultation were excluded due to incomplete data.

### Data Collection

Patient data was retrieved from clinical records and the e-hospital database. WHO-based case report forms were customized for local cryptococcosis surveillance and integrated into the REDCap platform (Research Electronic Data Capture, Vanderbilt University, US), hosted by AIIMS Rishikesh). Data fields included demographics, risk factors, clinical features, laboratory results, treatment, complications, and outcomes.

### Statistical Analysis

Data were analyzed using SPSS version 30. Categorical variables were expressed as frequencies and percentages; continuous variables as means ± SD or medians as appropriate. Between-group comparisons were made using Chi-square/Fisher’s exact test and t-test/Mann–Whitney U-test. Multivariate logistic regression was performed to determine predictors of mortality and critical illness. Statistical significance was set at P < 0.05.

### Ethical Considerations

The study was approved by the Institutional Ethics Committee of AIIMS Rishikesh (DHR REG NO: EC/NEW/INST/UA/0180 with approval number 352/IEC/IM/NF/2024). As the study was retrospective, informed consent was waived.

## Results

### Baseline demographic and geographic characteristics

A total of 33 patients diagnosed with cryptococcosis between January 2018 and December 2024 were included in the study (Table 1). The cohort comprised 26 males (78.8%) and 7 females (21.2%), with a median age of 44 years (range 19–66 years). Most cases (51.5%) were in the 30–45-year age group. Geographically, 11 patients (33.3%) were from Himalayan region and 22 (66.7%) from sub-Himalayan region (Figure 1 A-B).

**Fig. 1:**
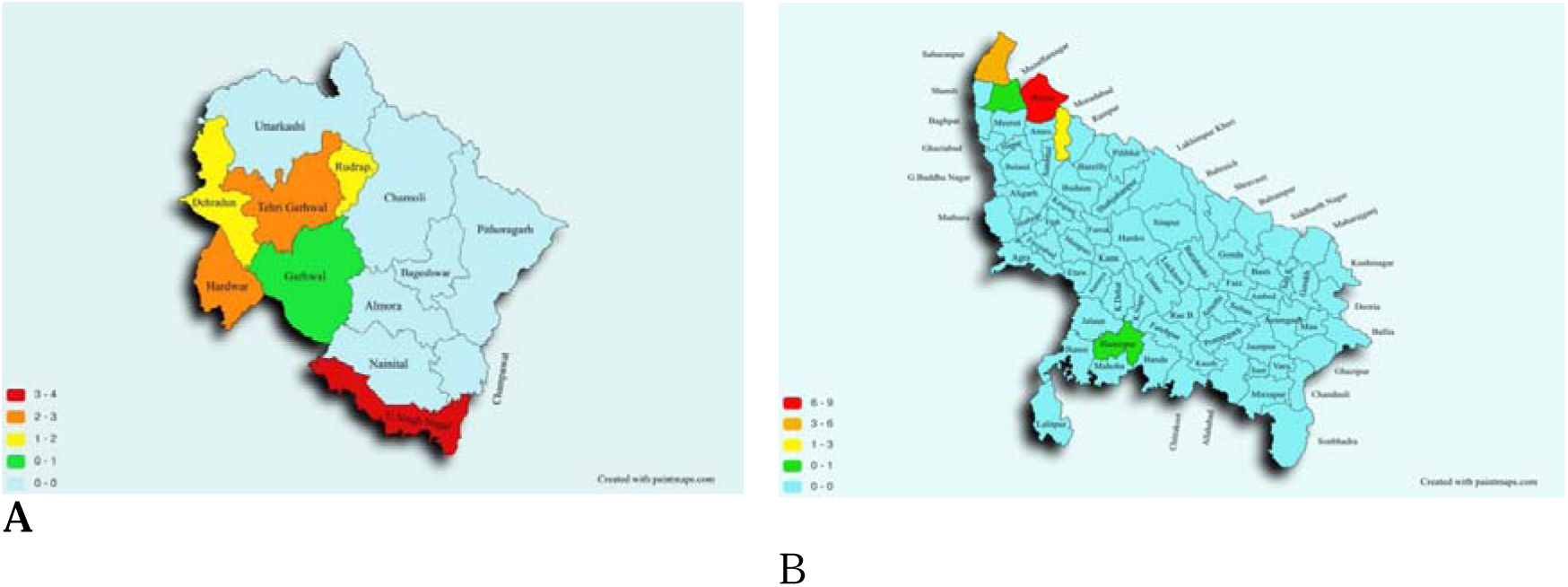
District Map of Indian state of Uttarakhand showing number of cases district wise (A) and of Uttar Pradesh showing number of cases district wise (B).

**Table 1:** Demography, comparison between HIV and Non-HIV sub-groups and Himalayan and Sub-Himalayan cryptococcosis patients.

|  |  |  |  |  |  |  |  |
| --- | --- | --- | --- | --- | --- | --- | --- |
| **Demography** |  |  |  |  |  |  |  |
| Gender |  |  |  |  |  |  |  |
| Male | 26<br>(78.8%) | 17<br>(77.3%) | 9<br>(81.8%) | 1.000 | 8 (72.7%) | 18<br>(81.8%) | 0.658 |
| Female | 7<br>(21.2%) | 5<br>(22.7%) | 2<br>(18.2%) |  | 3 (27.3%) | 4<br>(18.2%) |  |
| Age Group |  |  |  |  |  |  |  |
| 15–30 years | 4<br>(12.1%) | 2 (9.1%) | 2<br>(18.2%) | 0.586 | 1 (9.1%) | 3<br>(13.6%) | 1.000 |
| 30–45 years | 17<br>(51.5%) | 14<br>(63.6%) | 3<br>(27.3%) | 0.071 | 5 (45.5%) | 12<br>(54.5%) | 0.709 |
| 45–60 years | 10<br>(30.3%) | 5<br>(22.7%) | 5<br>(45.5%) | 0.240 | 4 (36.4%) | 6<br>(27.3%) | 0.693 |
| 60–75 years | 2 (6.1%) | 1 (4.5%) | 1 (9.1%) | 1.000 | 1 (9.1%) | 1 (4.5%) | 1.000 |
| **Risk Factors** |  |  |  |  |  |  |  |
| Reduced serum albumin | 21<br>(63.6%) | 13<br>(59.1%) | 8<br>(72.7%) | 0.703 | 8 (72.7%) | 13<br>(59.1%) | 0.703 |
| Immunosuppressive therapy | 3 (9.1%) | 2 (9.1%) | 1 (9.1%) | 1.000 | 1 (9.1%) | 2 (9.1%) | 1.000 |
| Diabetes mellitus | 2 (6.1%) | 2 (9.1%) | 0 (0.0%) | 0.542 | 2 (18.2%) | 0 (0.0%) | 0.105 |
| Organ transplant recipient | 1 (3.0%) | 0 (0.0%) | 1 (9.1%) | 0.333 | 0 (0.0%) | 1 (4.5%) | 1.000 |
| Chronic liver disease | 2 (6.1%) | 1 (4.5%) | 1 (9.1%) | 1.000 | 0 (0.0%) | 2 (9.1%) | 0.545 |
| **Clinical Manifestations** |  |  |  |  |  |  |  |
| Fever | 30<br>(90.9%) | 19<br>(86.4%) | 11<br>(100%) | 0.534 | 10 (90.9%) | 20<br>(90.9%) | 1.000 |
| Neck rigidity | 4<br>(12.1%) | 3<br>(13.6%) | 1 (9.1%) | 1.000 | 1 (9.1%) | 3<br>(13.6%) | 1.000 |
| Headache | 26<br>(78.8%) | 17<br>(77.3%) | 9<br>(81.8%) | 1.000 | 11 (100%) | 15<br>(68.2%) | 0.067 |
| Altered sensorium | 18<br>(54.5%) | 14<br>(63.6%) | 4<br>(36.4%) | 0.163 | 5 (45.5%) | 13<br>(59.1%) | 0.709 |
| Nausea/Vomiting | 18<br>(54.5%) | 12<br>(54.5%) | 6<br>(54.5%) | 1.000 | 5 (45.5%) | 13<br>(59.1%) | 0.709 |
| **Diagnostic Method** |  |  |  |  |  |  |  |
| India Ink positivity | 16<br>(48.5%) | 12<br>(54.5%) | 4<br>(36.4%) | 0.534 | 4 (36.4%) | 12<br>(54.5%) | 0.466 |
| CrAg (ICT) positivity | 9<br>(27.3%) | 4<br>(18.2%) | 5<br>(45.5%) | 0.121 | 4 (36.4%) | 5<br>(22.7%) | 0.430 |
| Culture positivity | 5<br>(15.2%) | 3<br>(13.6%) | 2<br>(18.2%) | 1.000 | 1 (9.1%) | 4<br>(18.2%) | 1.000 |
| **Treatment** |  |  |  |  |  |  |  |
| Amphotericin B | 30<br>(90.9%) | 20<br>(90.9%) | 10<br>(90.9%) | 1.000 | 8 (72.7%) | 22<br>(100%) | 0.030 |
| Flucytosine | 15<br>(45.5%) | 12<br>(54.5%) | 3<br>(27.3%) | 0.266 | 6 (54.5%) | 9<br>(40.9%) | 0.700 |
| Fluconazole | 22<br>(66.7%) | 17<br>(77.3%) | 5<br>(45.5%) | 0.117 | 6 (54.5%) | 16<br>(72.7%) | 0.417 |
| **Outcomes** |  |  |  |  |  |  |  |
| Discharged stable | 22<br>(66.7%) | 14<br>(63.6%) | 8<br>(72.7%) | 0.709 | 6 (54.5%) | 16<br>(72.7%) | 0.417 |
| LAMA | 4<br>(12.1%) | 3<br>(13.6%) | 1 (9.1%) | 1.000 | 2 (18.2%) | 2 (9.1%) | 0.591 |
| Death | 7<br>(21.2%) | 5<br>(22.7%) | 2<br>(18.2%) | 1.000 | 3 (27.3%) | 4<br>(18.2%) | 0.658 |
| **Morbidity Indicators** |  |  |  |  |  |  |  |
| HDU/ICU admission | 7<br>(21.2%) | 7<br>(31.8%) | 0 (0.0%) | 0.067 | 1 (9.1%) | 6<br>(27.3%) | 0.382 |
| Vasopressor requirement | 1 (3.0%) | 1 (4.5%) | 0 (0.0%) | 1.000 | 0 (0.0%) | 1 (4.5%) | 1.000 |
| Oxygen support | 2 (6.1%) | 2 (9.1%) | 0 (0.0%) | 0.542 | 0 (0.0%) | 2 (9.1%) | 0.545 |
| Septicemia | 5<br>(15.2%) | 4<br>(18.2%) | 1 (9.1%) | 0.643 | 1 (9.1%) | 4<br>(18.2%) | 0.648 |

### Clinical presentation and symptomatology

The most common presenting symptom was fever, seen in 30 patients (90.9%), followed by headache in 26 (78.8%), altered sensorium in 18 (54.5%), and nausea/vomiting in 18 (54.5%). Visual symptoms and muscle fatigue were reported in 24.2% and 15.2%, respectively. Neck rigidity was observed in only 12.1% of patients. Septicemia developed in 15.2% of patients, and a smaller subset required oxygen (6.1%) or vasopressor support (3%). One patient presented with papular skin lesions.

### Risk factors and comorbidities

HIV infection was the predominant comorbidity, affecting 22 patients (66.7%). Other notable risk factors included hypoalbuminemia (63.6%), use of glucocorticoids or immunosuppressive therapy (9.1%), diabetes mellitus (6.1%), chronic liver disease (6.1%), and organ transplantation (3%).

### Diagnostic methods

Among 33 patients, India ink staining was positive in 16 patients (48.5%), cryptococcal antigen positivity (ICT) was found in 10 patients (30.3%), fungal growth on culture was noted in 6 cases (15.2%), and histopathology was the diagnostic test in 1 patient (3.0%) who presented with skin lesions.

### Therapeutic management

Amphotericin B deoxycholate was administered to 30 patients (90.9%) at a dose of 0.7–1.0 mg/kg/day, with a mean duration of 12.3 ± 4.1 days. Flucytosine was used in 15 patients (45.5%) at 100 mg/kg/day in divided doses, typically for 11.6 ± 3.5 days. Fluconazole was prescribed to 22 patients (66.7%) at 400–800 mg/day, mainly during the consolidation phase, with a mean maintenance duration of 30.4 ± 8.2 days. Treatment regimens were chosen based on clinical severity, renal function, and drug availability, following IDSA and WHO guidelines. The average total duration of hospitalization was 17.06 days (SD ± 9.94). Seven patients (21.2%) required high-dependency or intensive care support, with a mean ICU stay of 7.43 days (SD ± 4.86).

### Outcomes and mortality

Of the 33 patients, 22 (66.7%) were discharged stable, 4 (12.1%) left against medical advice, and 7 (21.2%) died during hospitalization. After 1 year of follow-up data for 26 patients, 16 of these patients were alive and 10 had died. Combining these 10 post-discharge deaths with the 7 in-hospital deaths, the overall 1-year mortality was 17 of 33 patients (51.5%). Amphotericin B administration showed a protective association (OR 0.4), while uncontrolled diabetes, ICU admission, and altered sensorium had negative odds (Figure 2).

**Fig 2:**
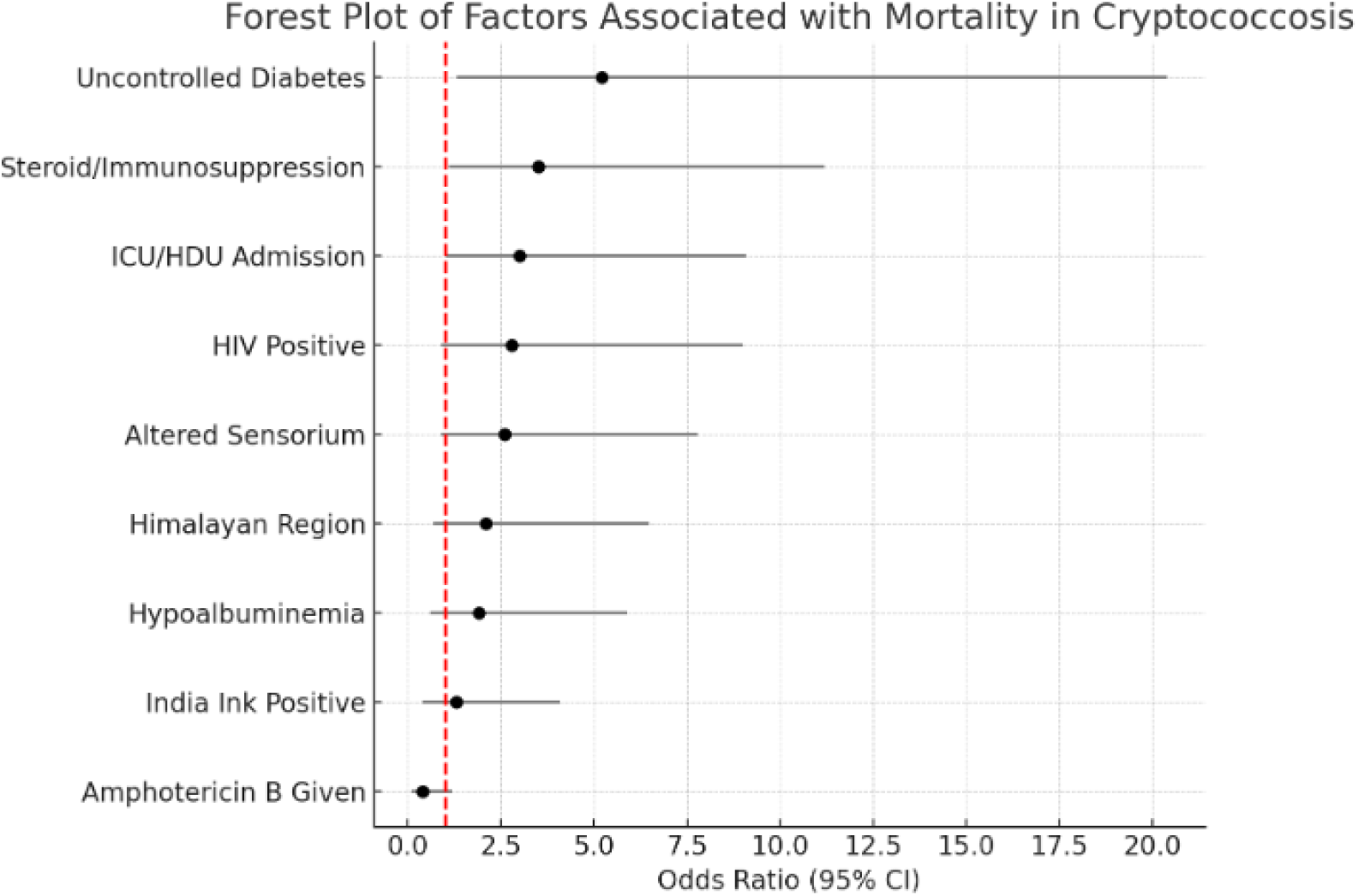
Forest plot showing logistic regression analysis of risk factors associated with mortality in patients with cryptococcosis. The red dashed line represents an odds ratio (OR) of 1. Variables with OR > 1 indicate increased odds of mortality; OR < 1 indicates protective effect. Amphotericin B administration showed a protective association (OR 0.4), while uncontrolled diabetes, ICU admission, and altered sensorium were notable risk factors.

### Predictors of adverse outcomes

Uncontrolled diabetes mellitus (HbA1c > 6.5%) was the only variable significantly associated with in-hospital mortality (p=0.019). The presence of HIV infection (p=0.035) and prior corticosteroid or immunosuppressive use (p=0.043) were significantly associated with ICU/HDU admission. No statistically significant associations with death were found with old age, gender, or immune status (Table 2).

**Table 2:** Predictors of Mortality – Logistic Regression Analysis

| Variable | Odds Ratio (OR) | 95% CI | P value |
| --- | --- | --- | --- |
| Age | 1.057 | 0.912 – 1.226 | 0.463 |
| Male gender | 0.351 | 0.017 – 7.028 | 0.493 |
| Immunocompromised status | 0.711 | 0.048 – 10.622 | 0.805 |

### Subgroup analysis: HIV vs non-HIV

Comparison between HIV-infected (n=22) and non-infected (n=11) patients showed no statistically significant differences in any clinical or laboratory parameter (Table 1). However, certain trends were notable: altered sensorium, diabetes, need for ICU admission, septicemia, and mortality were more common among the HIV group. Non-HIV patients had higher incidences of headache, reduced serum albumin, and chronic liver disease. Amphotericin B was uniformly administered, while flucytosine and fluconazole were more frequently used in HIV patients, albeit without statistical significance.

### Subgroup analysis: Himalayan vs sub-Himalayan cohorts

No statistically significant difference was observed in the clinical features or outcomes between Himalayan (n=11) and sub-Himalayan (n=22) patients (Table 1). However, neck rigidity, altered sensorium, chronic liver disease, and ICU admission were more common in the sub-Himalayan group. Headache, hypoalbuminemia, diabetes, and mortality were more frequently observed in the Himalayan group. Notably, amphotericin B usage was significantly lower in the Himalayan group (72.7% vs 100%, p=0.0302).

## Discussion

Cryptococcosis poses a persistent public health challenge, especially in low- and middle-income countries where immunocompromised populations are expanding due to the HIV epidemic and rising prevalence of comorbid conditions. In this retrospective study spanning seven years at a tertiary care center (AIIMS Rishikesh) in Northern India, we evaluated the clinical and epidemiological landscape of microbiologically confirmed cryptococcosis cases. Our findings not only underscore established risk associations but also highlight evolving patterns in disease demographics, diagnostics, and treatment outcomes—particularly within the underserved Himalayan belt.

One of the key demographic insights from our cohort is the predominance of middle-aged males, with most cases occurring in the 30–45-year age bracket. This reflects the demographic overlap with HIV-infected populations, which remain disproportionately affected by cryptococcal infections globally [1]. However, nearly one-third of our patients were HIV-negative, pointing to a growing spectrum of hosts at risk. This includes individuals with chronic metabolic diseases such as diabetes mellitus, those on prolonged corticosteroid therapy, and patients with hypoalbuminemia—conditions known to impair immune defense mechanisms against fungal pathogens [10] [11]. Diabetes mellitus impairs host defense through multiple mechanisms, including reduced chemotaxis, impaired neutrophil phagocytosis, and defective cytokine production. Hyperglycemia also promotes fungal growth and disrupts the integrity of the blood-brain barrier, facilitating CNS invasion [12] [13]. In a large study from China, diabetic patients with cryptococcosis were shown to have higher mortality and longer hospital stays than non-diabetics [14]. Chronic corticosteroid use, often prescribed for autoimmune diseases or organ transplantation, diminishes cellular immunity by inducing lymphocyte apoptosis and suppressing Th1-type immune responses. This dampens macrophage activity and impairs the body’s ability to mount a granulomatous response to fungal pathogens like Cryptococcus neoformans. Corticosteroids also reduce antigen presentation and blunt inflammatory responses, further masking the clinical presentation and delaying diagnosis [15]. Hypoalbuminemia, commonly seen in chronic liver disease, malnutrition, and critically ill patients, serves as a marker of poor systemic resilience and impaired immune competence. Albumin plays a role in maintaining endothelial integrity and serves as a negative acute-phase reactant. Low serum albumin levels are independently associated with increased risk of invasive fungal infections and poor outcomes, including in cryptococcal meningitis. Studies have shown that hypoalbuminemia correlates with increased blood-brain barrier permeability, which may facilitate fungal dissemination to the CNS [16].

Neurological involvement was the hallmark presentation in our patients, with fever and headache observed in over 75% of cases. These features are consistent with cryptococcal meningoencephalitis, a condition that often progresses insidiously and may be mistaken for tuberculous meningitis in endemic regions like India [17] [18]. Surprisingly, classical meningeal signs such as neck rigidity were infrequent, reported in just over 10% of patients. This highlights the clinical ambiguity of cryptococcal CNS infection, necessitating a high index of suspicion and routine use of lumbar puncture for early diagnosis [19].

Diagnostic modalities relied heavily on India ink microscopy, which, is inexpensive and rapid. Cryptococcal antigen (CrAg) detection, though more sensitive, was positive in 30.3% of our cases. Expanding access to lateral flow assays and integrating CrAg screening into standard protocols for high-risk groups could significantly reduce diagnostic delays.

Our study reinforces the continued relevance of amphotericin B-based regimens, administered in over 90% of patients. Flucytosine and fluconazole were employed in fewer cases, often dictated by availability and renal tolerance.

Outcome data reveal an in-hospital mortality rate of 21.2%, which rose to nearly 30% when post-discharge deaths were included. These figures are consistent with global mortality estimates for cryptococcal meningitis but are concerning given advancements in antifungal therapy [20] [10]. Notably, uncontrolled diabetes mellitus (HbA1c >6.5%) was the only variable significantly associated with in-hospital death (p = 0.019). Hyperglycemia is known to impair neutrophil function and macrophage activity, predisposing individuals to invasive fungal infections and worsening prognosis [9] [21].

The forest plot (Figure 5) offers valuable visual insight into factors influencing patient survival. Variables such as ICU admission, altered mental status, and diabetes showed positive associations with mortality risk (OR > 1), while amphotericin B use had a protective effect (OR < 1) [22].

Subgroup analysis by HIV status did not yield statistically significant differences in clinical manifestations or outcomes, but trends were notable. HIV-positive patients more frequently required intensive care, likely due to advanced disease and co-infections. In contrast, HIV-negative patients showed higher rates of liver disease and hypoalbuminemia, supporting the theory that nutritional and metabolic immunosuppression plays a growing role in disease susceptibility. These observations suggest that clinicians should consider cryptococcosis in a broader spectrum of immunocompromised hosts—not just those with HIV.

A second layer of subgroup analysis compared Himalayan and sub-Himalayan cohorts. Himalayan patients presented late in the disease course, for this training local physicians to recognize atypical fungal presentations and initiating empirical antifungal therapy based on clinical suspicion may mitigate these regional inequities. Structured training and clinical mentorship of rural healthcare workers improved diagnostic accuracy, treatment adherence, and patient outcomes in resource-limited settings—outcomes that are directly applicable to diseases like cryptococcosis, where early suspicion and treatment are crucial.

The strength of this study lies in its real-world relevance. By profiling cryptococcosis in a diverse Indian population, including underrepresented Himalayan communities, we underscore critical gaps in diagnosis, treatment access, and outcomes. Moreover, prospective multicenter studies with molecular surveillance are needed to track evolving pathogen profiles and resistance trends in the Indian subcontinent.

Study limitations

This study has certain limitations. First, it is a retrospective analysis, and thus subject to documentation bias and missing data. Second, follow-up information beyond discharge was not available for all patients, limiting the ability to assess long-term outcomes or relapse. Third, sample size was relatively small, especially for subgroup analyses such as transplant recipients or patients with autoimmune diseases. Lastly, drug susceptibility testing of *Cryptococcus* species and molecular typing were not performed due to resource constraints.

## Conclusions

Cryptococcosis remains a major opportunistic fungal infection in Northern India, predominantly affecting people living with HIV but increasingly recognized in individuals with diabetes, chronic illnesses, or immunosuppressive therapy. This study highlights the high burden of cryptococcal meningitis, the importance of early diagnosis through antigen testing, and the need for timely access to amphotericin-based therapy. Mortality remains substantial, especially among patients with uncontrolled diabetes or limited access to optimal antifungal regimens. Strengthening decentralized diagnostic capabilities, ensuring antifungal availability in remote settings, and recognizing at-risk non-HIV populations are crucial to improving clinical outcomes in cryptococcal disease.

## DATA SHARING

It will be made available to others as required upon requesting the corresponding author.

## Data Availability

All data produced in the present study are available upon reasonable request to the authors.

## ACKNOWLEDGMENT

None

